# The allometric propagation of COVID-19 is explained by human travel

**DOI:** 10.1101/2021.04.08.21255169

**Authors:** Rohisha Tuladhar, Paolo Grigolini, Fidel Santamaria

## Abstract

We analyzed the number of cumulative positive cases of COVID-19 as a function of time in countries around the World. We tracked the increase in cases from the onset of the pandemic in each region for up to 150 days. We found that in 81 out of 146 regions the trajectory was described with a power-law function for up to 30 days. We also detected scale-free properties in the majority of sub-regions in Australia, Canada, China, and the United States (US). We developed an allometric model that was capable of fitting the initial phase of the pandemic and was the best predictor for the propagation of the illness for up to 100 days. We then determined that the power-law COVID-19 exponent correlated with measurements of human mobility. The COVID-19 exponent correlated with the magnitude of air passengers per country. This correlation persisted when we analyzed the number of air passengers per US states, and even per US metropolitan areas. Furthermore, the COVID-19 exponent correlated with the number of vehicle miles travelled in the US. Together, air and vehicular travel explained 70 % of the variability of the COVID-19 exponent. Taken together, our results suggest that the scale-free propagation of the virus is present at multiple geographical scales and is correlated with human mobility. We conclude that models of disease transmission should integrate scale-free dynamics as part of the modeling strategy and not only as an emergent phenomenological property.

## Introduction

There is an increasing number of studies showing that the number of positive cases of COVID-19 have power-law dynamics (1-9). If indeed, the propagation of the illness is governed by scale-free processes and is not only an emergent dynamic from complex interactions that assume exponential growth then, modeling strategies might have to be revised. However, except for a few cases (3), these studies have focused on a few countries or regions. Thus, it is important to understand how prevalent power-law propagation is around the World. It is also important to understand if a power-law spread of the illness measured at a national level is reflected in its sub-regions. Furthermore, since policies on restricting the spread of the virus have included reducing mobility, it is necessary to find how that affects the power-law properties of the propagation of COVID-19 (10).

Traditional modeling approaches to illness propagations, such as COVID-19, assume an exponential growth (11-16). However, increasing evidence shows that human dynamics have power-law properties (17-20). In fact, recent work has shown that multiple measurements of human activity follow allometric properties such as *w* ∝ *N*^*α*^, where N is the number of individuals and w a metric of activity (21). Since most of the world population now lives in urban areas (22) it is important to consider how such urban human dynamics affects the spread of illnesses. Furthermore, since asymptomatic transmission is a prevalent feature of SARS-Cov-2 (as the virus that causes COVID-19) (23) then the virus will propagate in a population following the dynamics of the healthy population, as opposed to illnesses in which the probability of infections is highest when individuals are symptomatic (24). Therefore, the spread of COVID-19 could be influenced by allometric properties observed in urban areas.

In this project, our aim was to understand the spread of the virus at the beginning of the pandemic. For this, we analyzed the total positive cases of COVID-19 in all countries and regions in the World. We also analyzed sub-regional behavior in four countries in three continents. Furthermore, we studied the spread of the virus in the United States (US) at the national, state, and urban levels. We found that the majority of countries followed a power-law in the cumulative number of positive cases of COVID-19 for at least 30 days. The majority of the sub-regions of the countries we studied, including 33 of the 50 US states, followed power-law dynamics. We built an allometric growth model that was able to replicate the power-law increase in COVID-19 cases. More interestingly, the model was also capable of capturing the dynamics of the initial days of the pandemic, which did not follow power-law increases. We tested the model’s predictive power by extrapolating its values up to 100 days and compared to actual data. We show that the allometric growth model is better than a simple power-law or exponential fits. We end the study by showing that the value of the COVID-19 power-law exponent is correlated with air travel at the world, national, and metropolitan scales. For the US we show that 70 % of the variance of the value of the power-law exponent is explained by air and vehicular travel. Overall, our work shows that power-law behavior of the spread of COVID-19 is observed across scales, it is related to human mobility, and that allometric models are a good strategy to increase the predictive power of computational studies.

## Results

### Power-law propagation of COVID-19 at the global and regional scales

We studied the number of cumulative test positive cases of COVID-19 (*I*) in 187 countries and territories from 22 January 2020 to 23 August 2020. We wanted to obtain a date that acted as reference for a period of sustained increase. For this purpose, for each region, we started our analysis after *I* ≥ (*I*_*Th*_ = 100). We did this to avoid plateaus and sudden changes in cases reported in the first days of the pandemic. Then, we calculated the ratio between 21 days (*R* = *I*(*t* + 21)/*I*(*t*)) and we selected the day (*I*_*o*_) when *R*_2_ ≥ 2 for the first time. As a result, we obtained a list of 146 regions. The other 41 regions included countries with small populations; regions that had a spread of the pandemic slower than *R*_2_; or than when they reached that threshold did not have enough data points for analysis (at least 35 days). We then fitted the cumulative cases per day to a power-law, *I* = *t*^*α*^ + *b*, where *α* is the COVID-19 exponent; or an exponential function, *I* = *A exp*(*t*/*τ*) + *d*, where τ is the time constant. In order to compare the fits, we fitted all the traces over a range that started *F*_*s*_ = 5 days after *I*_*o*_ and lasted 30 days (*F*_*range*_), except for Australia (*F*_*range*_=15) and China (*F*_*s*_= 3 and *F*_*range*_ = 10) because of their early strict social mobility policies. The mean value of cumulative cases on *I*_*o*_ was 124.31 ± 5.44 SEM cases. We calculated the root mean square error (RMSE) and the r-square values for each fit. We determined the goodness of fit by comparing the RMSE of the power-law and exponential fits. The fit with the lowest RMSE had to have an r-square larger than 0.97. This analysis showed 81regions were better fit by a power-law, 31 were better explained by an exponential fit, and 34 by neither (see Supplementary Materials). We plotted the trajectories of all regions fitted by a power-law for 100 days after their respective *I*_*o*_ (107 of the 146 areas had enough data points). This showed that the propagation followed a similar trajectory (Figure 1, the Supplementary Materials contain the table form data). The average value of the COVID-19 exponent was 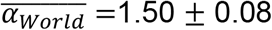 SEM with an inter-quantile range of 0.99. The propagation of COVID-19 seems to follow a different trajectory than previous pandemics. For example, we analyzed the total positive cases during the Ebola epidemic in West Africa and influenza deaths during the 1918 pandemic in Philadelphia and found that both follow exponential fits (25) (Supplementary Materials Figure S1). Thus, our analysis suggested that most of the regions in the world experienced a power-law increase in the propagation of COVID-19.

**Figure 1.**
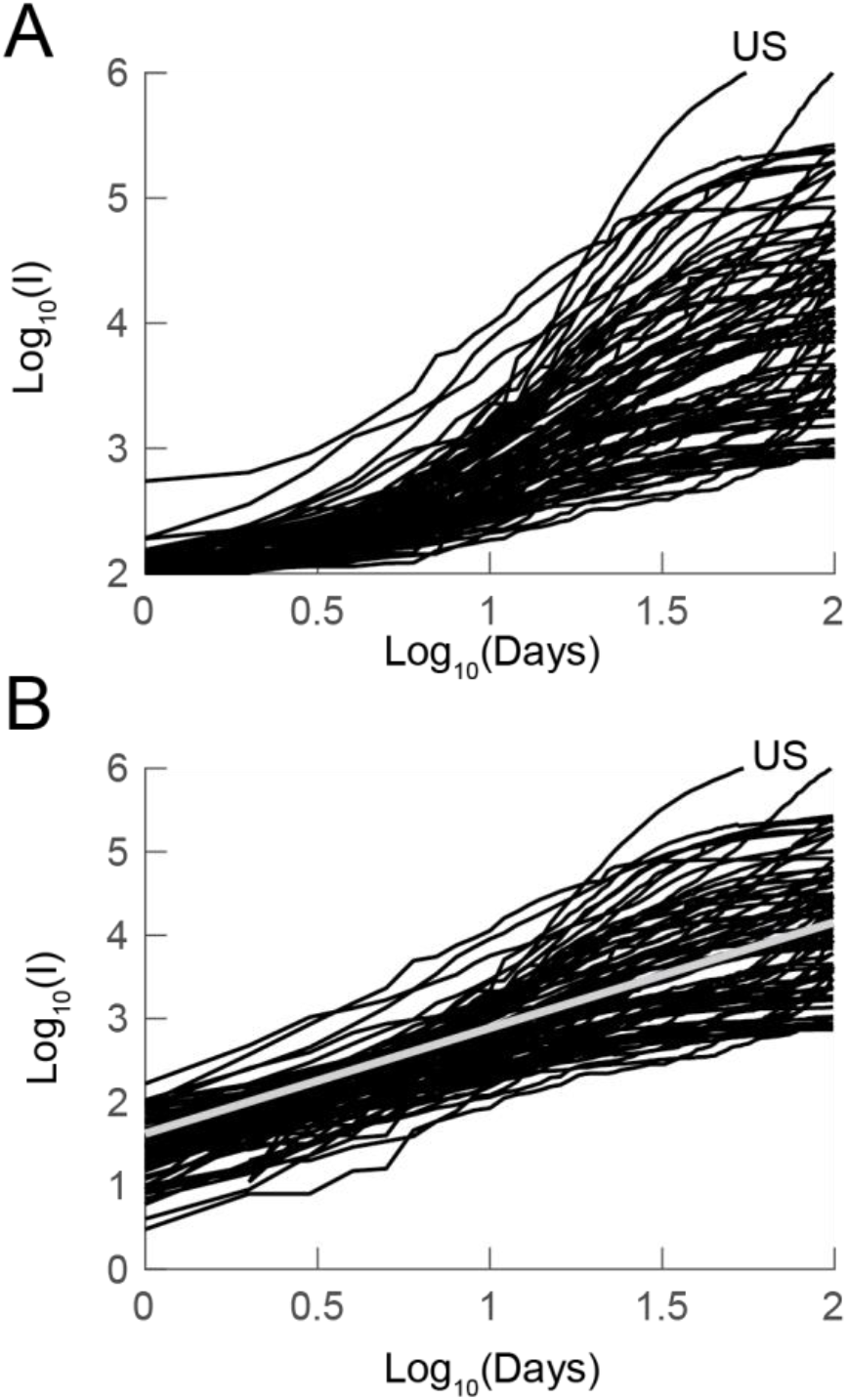
Power-law propagation of COVID-19 across the World. A) cumulative positive cases reported for 81 countries and regions over time. B) same data as in A but the trajectories were referenced to their first value. The gray line is the fit to the mean of the trajectories resulting in *Log*10(*I*) = 1.27 ∗ *t* + 1.62. *I* is the cumulative number of cases for each country. See text for details.

We were interested in understanding if the propagation of COVID-19 followed power-law dynamics in sub-regions within countries. For this, we chose Australia, Canada, China, and the Unites States (US). In the case of Australia, our algorithm determined that the propagation of COVID-19 followed an exponential function. Never the less, we studied the propagation of the pandemic in all Australian states. In order to detect the onset of activity in a smaller population we used an *I*_*Th*_ = 20 (Figure 2a). Our regional analysis shows that 5 of the 8 Australian states had a power-law propagation with a mean of 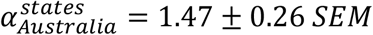. The states that followed power-law dynamics included most of the Australian population. However, the most populous state, New South Wales, followed exponential growth. Another aspect that did not allow the power-law dynamics be reflected nationally was that the start of the epidemic in each state (day of first reported case) took place over 18 days. Thus, while the propagation of the illness appeared exponential at the national level the majority of the country showed power-law behavior.

**Figure 2.**
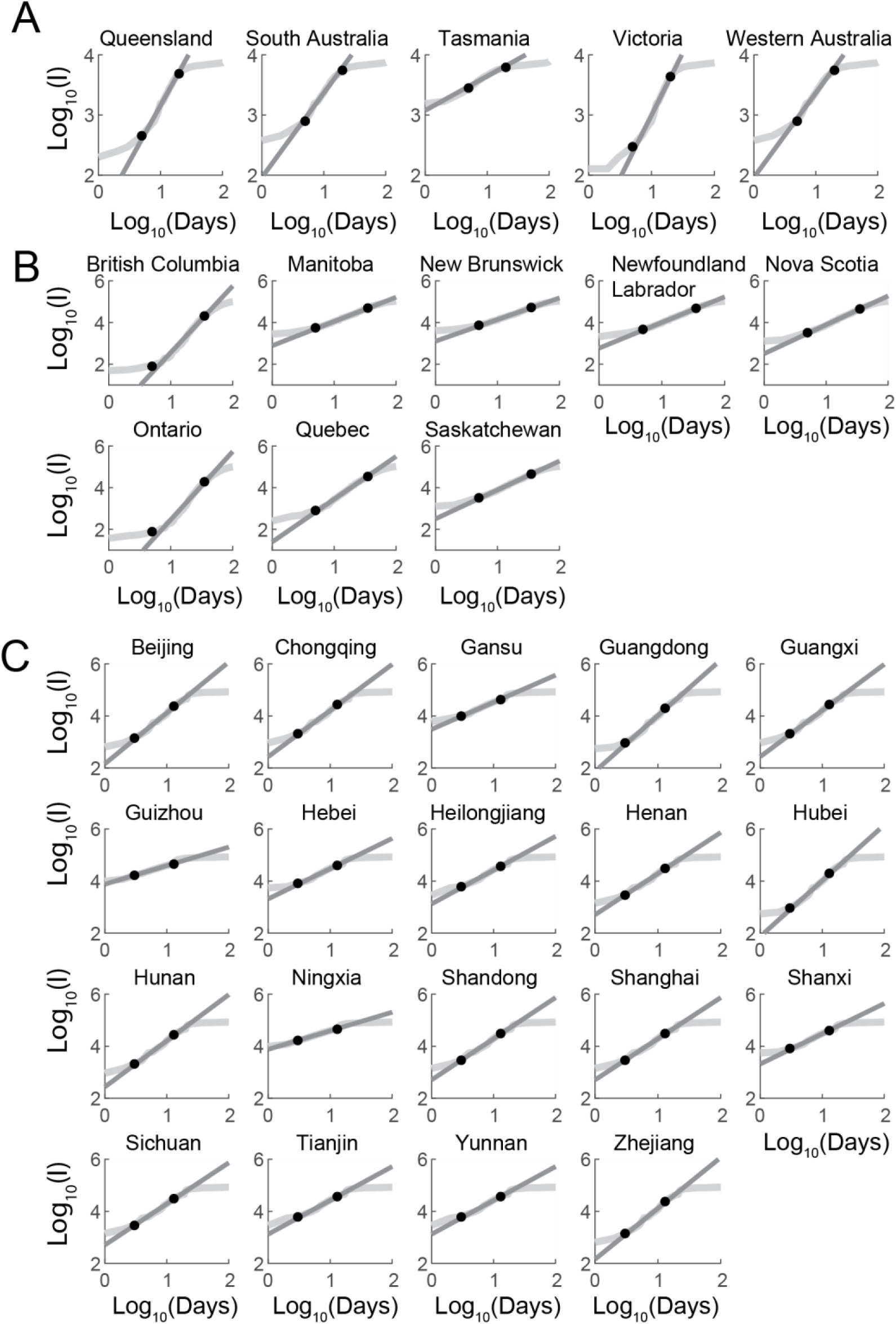
The sub-regions of Australia, Canada, and Chine follow power-law increase in cases of COVID-19. A) Regions in Australia that has power-law behavior. The black dots indicate the first and last point used for the fitting, all ranges were the same for each sub-region. The gray straight line is the plot of the fit. B) and C) same analysis for the provinces of Canada and China that followed power-law behavior.

The same analysis in Canada showed that 8 out of 14 provinces and territories followed power-law propagation with an average of 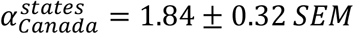. In this case, the national propagation of COVID-19 also followed a power-law increase with a value of *α*_*canada*_ = 2.57 ± 0.08 *CI*95. In this case, the national result was influenced by the faster propagation of COVID-19 in more populous provinces, with British Columbia 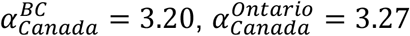, and 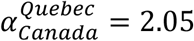 (Figure 2b). In China, 19 of the 30 provinces were also described with a power-law. The average propagation of 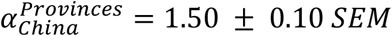 was again slower than for the entire country (*α*_*China*_ = 2.14 ± 0.17 *CI*95), which was dominated by Hubei, where the virus originated 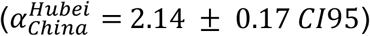 (Figure 2c). Finally, for the US, we used *I*_*Th*_ = 10, *F*_*s*_ = 5, and *F*_*Range*_ = 20. This analysis showed that 33 of 50 states had power-law dynamics 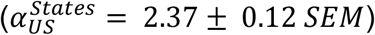. Again, the analysis for the entire country shows a faster propagation (*α*_*US*_ = 3.98 ± 0.21 *CI*95) (Figure 3). Overall, our regional analysis shows that COVID-19 propagated following power-law dynamics in the early stages of the pandemic.

**Figure 3.**
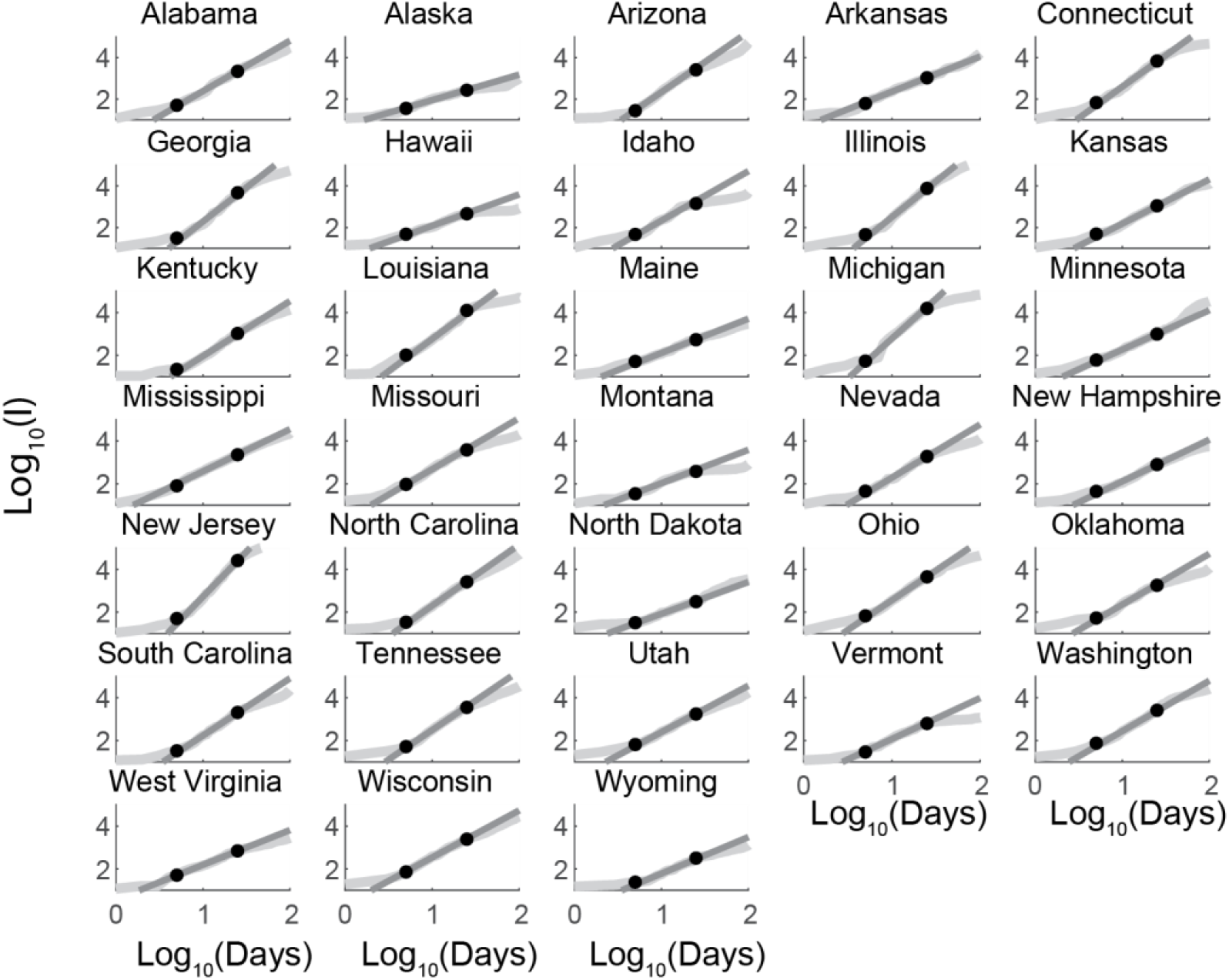
The propagation of COVID-19 in the US follows a power-law behavior in most states. The cumulative number of cases (I) were analyzed after surpassing 100 cases in each state. In all cases the power-law function was fitted 5 days after the reference date for 20 days. See text for details.

### An allometric model predicts the spread of cases

The propagation of COVID-19 seems to have been driven by asymptomatic infected individuals. This is different from other illnesses in which patients are infectious when showing symptoms. Thus, the modeling approach should be based on the behavior of healthy populations. Increasing information shows that the metrics of metropolitan community activity have a power-law, or allometric, scaling with the size of the population (21). We assumed that human activity, for example travel, shows such behavior and thus the activity of the infected population could be described by:

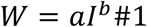

As postulated by others (see eq 4.65 and 4.67 in (26)), the state of resources used by a population can be described by:

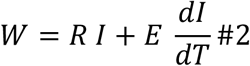

Re-arranging and renaming variables we obtained:

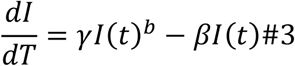

Where *γ* is the classical rate of infections and, in our simplified model, *β* is the number of recovered or diseased. This model is equivalent to one in which the rate of infection depends on time:

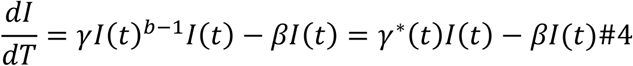

In the Methods section, we show that the solution to equation 3 has a temporal regime in which the model has power-law behavior, *I ∝ t*^*α*^, and the relation of b with *α* is:

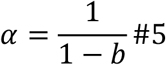

With *b* < 1.

We fitted the solution of our allometric model using the same parameters used to fit the power-law function. We used the value of *α* that we calculated before and allowed the fitting algorithm to get values for *γ* and *β*. While fitting the data from the start of the spread was not our objective, we compared the values predicted by the fits to the number of cases at *I*_0_. This shows that in the allometric fit deviated −52 %, the power-law −73 % and the exponential 181 % (see examples in Figure 4a and 4b). These results suggested that the allometric model could better fit the behavior of cases in the initial days of the pandemic. In order to compare the predictive power of the power-law, exponential, and allometric fits we calculated the percentage difference between the models and real data for up to 100 days after the last day used for the fit (only 20 regions had enough data points for this analysis). This analysis shows that the allometric fit is the best model to predict the propagation of COVID-19 (Figure 4a, right). The same analysis for US states also shows that the allometric model provides reliable predictions of the evolution of the spread (Figure 4b). Overall, our allometric model is a parsimonious framework to model the initial spread of the pandemic and provides strong predictive power.

**Figure 4.**
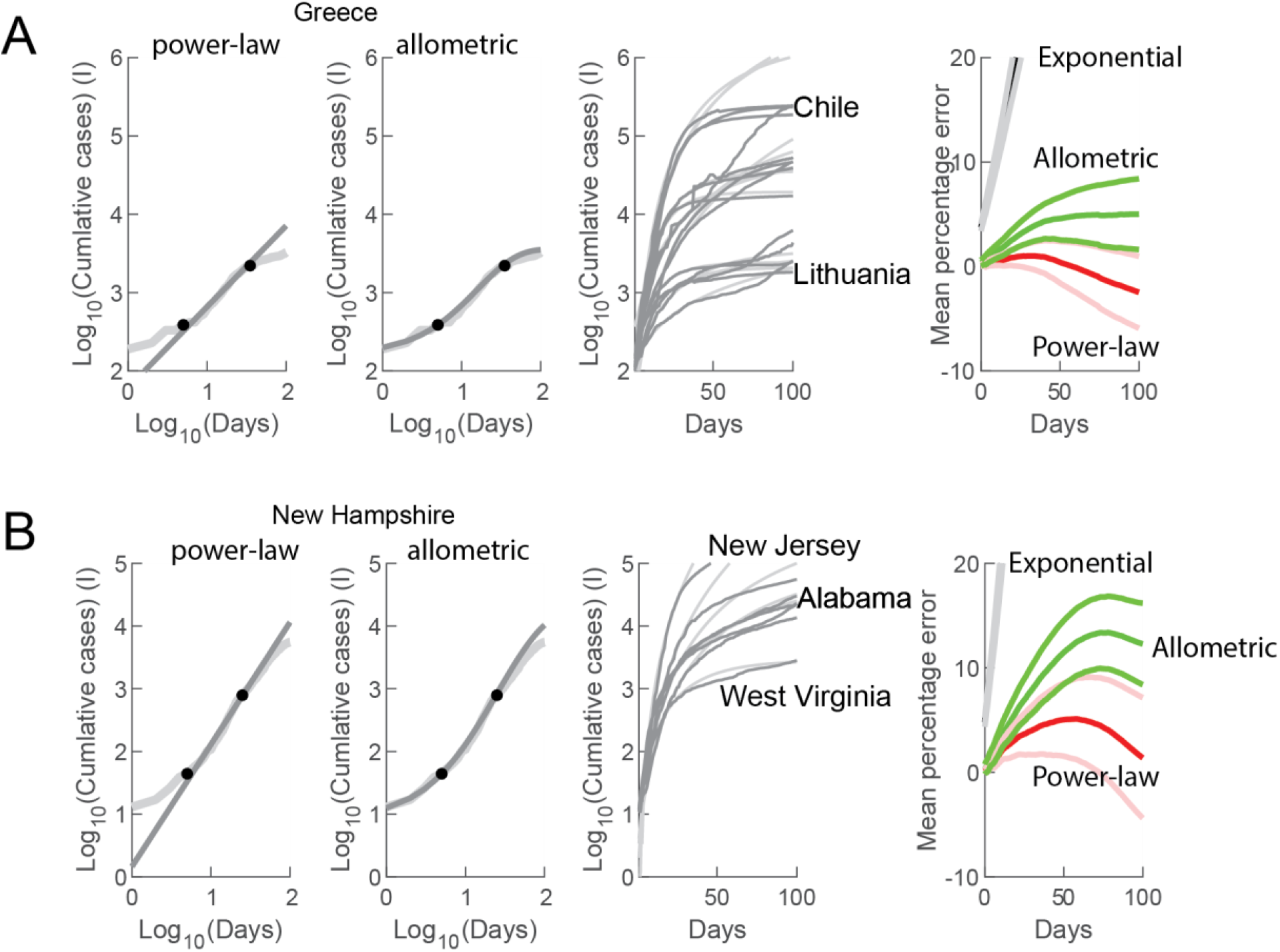
The allometric model predicts the behavior of the propagation COVID-19. A) Left: example of fitting a power-law and allometric function to the same date of cumulative cases (I) of COVID-19 for Greece. Middle: Examples of cumulative cases and allometric predictions for up to 100 days after the date of the last point used to fit the curves. Right: The mean percentage error for 20 countries over 100 days after the last point used to fit the exponential (black), power-law (red), and allometric (green) functions. The lighter shade curves correspond to the 95 % confidence intervals. B) same as in A applied to 33 states that follow power-law dynamics in the United States.

### Human travel is correlated with propagation of COVID-19 from countries to cities

Lockdown measures enacted to reduce the spread of COVID-19 included the reduction of human mobility. In this context, we wanted to understand how travel affects the value of *α*. As such, we analyzed the relationship between the number of air passengers per country against the value of *α*. We found that the magnitude (Log_10_) of the number of passengers strongly correlated to the value of *α*, with 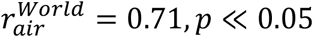 (Figure 5a). We repeated this analysis for the number of air passengers in the 33 US states that were described with a power-law. Here again we found a strong correlation, 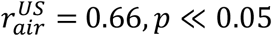 (Figure 5b). We also analyzed the countries and US states that were better described by an exponential fit. In this case, we again found a significant correlation between the value of the calculated time constant and the magnitude of air travel (Supplementary Figure S2). This suggested that the processes represented by a time constant could be also part of a slower power-law process. Furthermore, we analyzed the spread of COVID-19 in 30 metropolitan areas that corresponded to the largest airports by number of passengers in the US. From these, 23 metropolitan regions serviced by those airports showed a power-law increase and there was a correlation coefficient of 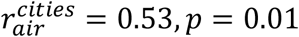 (Figure 5c). Together, our analysis suggests a strong effect of air travel magnitude in the power-law propagation of COVID-19.

**Figure 5.**
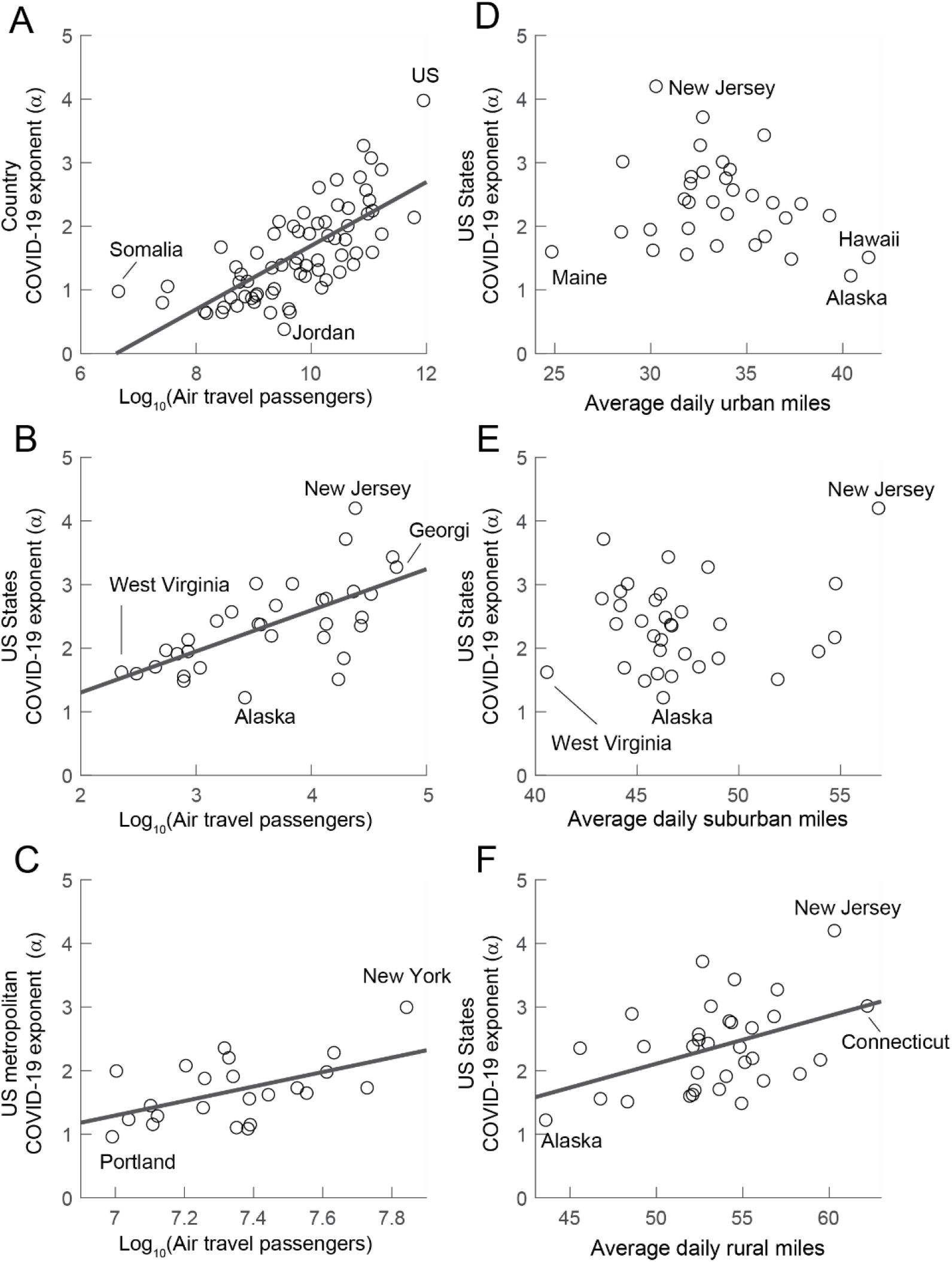
The power-law COVID-19 exponent (*α*) correlates with human travel. A) scatter plot of the value of *α* against the scale of total number of passengers enplaned in 2017. B) and C) same as in A but for total passengers per state in the US (B) and for metropolitan areas served by the major airports in the US. D-F) as in A-D for the average miles traveled by vehicle in urban (D), suburban (E), and rural (F) areas. Lines were plotted when the correlations were significant. See text for details.

The previous analysis suggest that air travel affects the dynamics of the propagation of the virus. However, this could be due to long-distance movement and not directly related to air travel. In order to compare the propagation of the virus to other forms of travel we studied vehicle miles traveled in the US. We obtained the data for urban, suburban, and rural travel per state. Our results show that there is no correlation between the value of *α* per state against the corresponding average urban miles 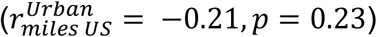 and suburban miles travelled (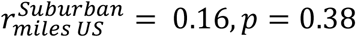,Figures 5d and 5e). However, there is a positive correlation with rural miles traveled (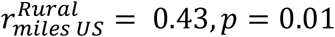, Figure 5f). There was no correlation between the number of air passengers per state and any of the three measurements of miles traveled per state (although the urban miles were almost significant, see Supplementary Materials), suggesting that the two modes of transport contribute independently to the propagation of the virus. Finally, we constructed linear models for all the combinations of air passenger and miles traveled by state against their respective value of *α*. This showed that air passengers accounted for 43 % of the observed variance of *α* but when combined with the three measurements of miles traveled by vehicle the model accounts for 70 % of the variance observed in the value of *α* (Figure 6).

**Figure 6.**
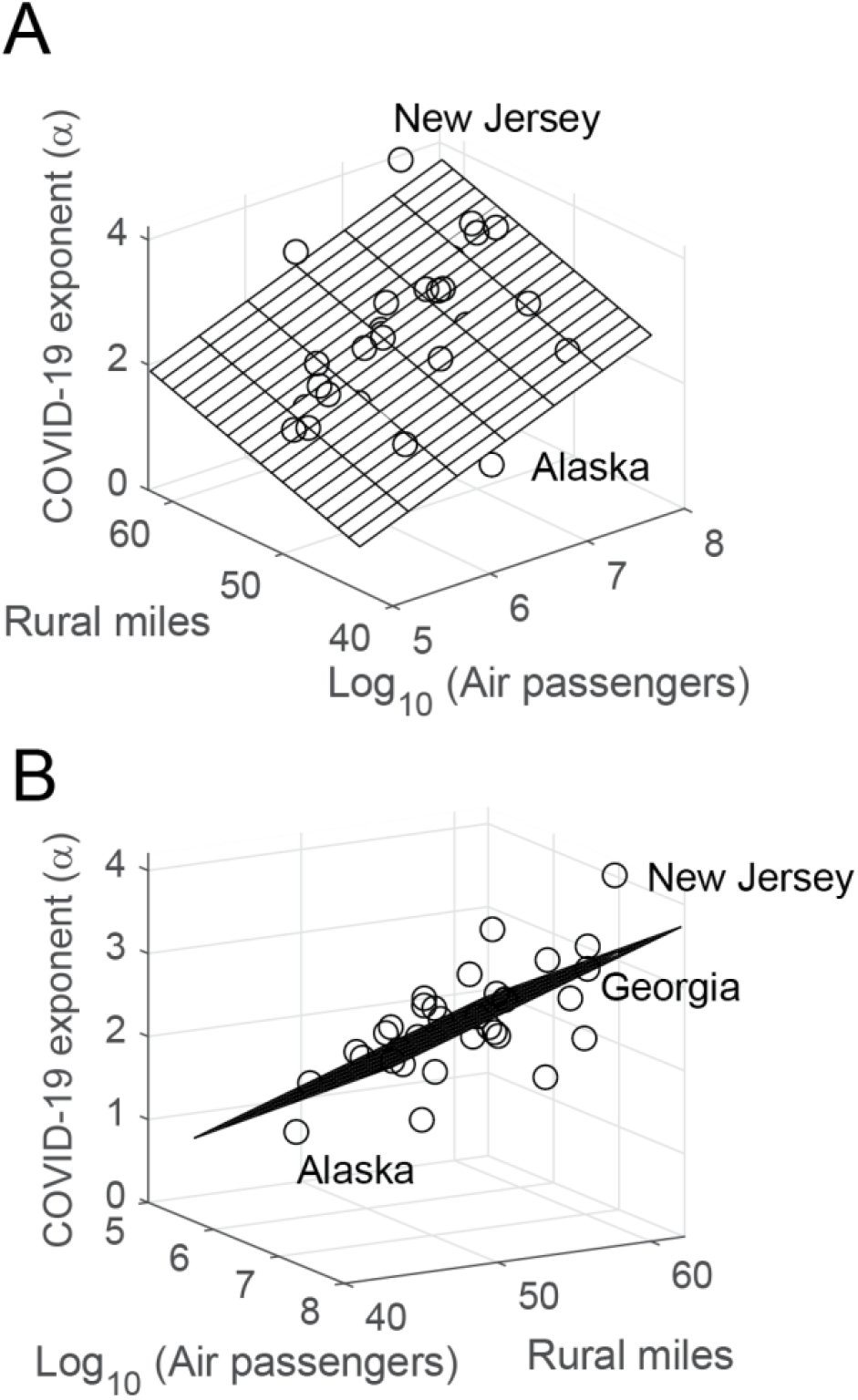
The variance observed in the power-law COVID-19 exponent (*α*) in the US is explained by air and vehicle travel. A) Rendering of the values of *α* versus number of air passengers and rural miles traveled per US state. The linear model (grid) explains 57 % of the variance. A model including urban and suburban miles explains 70 %. B) a different view angle of the same data. See text for details.

## Discussion

The power-law propagation of COVID-19 was reported recent in the pandemic (9, 27). In our current work, we found that the total positive cases of COVID-19 infection follows a power-law increase in the majority of the regions of the World as well as at the regional level in Australia, Canada, China, and the US.

The emergence of power-law propagation of the virus could be due to different time constants of the propagation that together appear as a power-law or due to the effect of lockdown measures (10). However, we showed that power-law behavior is present in sub-regions of geographically large countries (Australia and Canada) or with big populations (China and US). We went further to show that metropolitan regions served by the busiest airports also showed power-law dynamics. This multi-scale property suggests that the propagation of the virus is following scale-free human dynamics properties (28).

The power-law infection dynamics could also be comparable to the experimentally observed power-law dispersal of bank note trajectories (28) which was used as a proxy for human travel on all geographical length scales. Other previous work (29, 30) showed that the spread of SARS 2003 can be reproduced by a model that considers nearly the entire civil aviation network; however, this model could only describe global trends but not local spatial scales. Other work has found an association between mobility patterns and COVID-19 transmission in the US (31). Their analysis showed that mobility patterns were strongly correlated with decreased COVID-19 case growth rates for the most affected counties in the US. An early analysis of COVID-19 propagation show the importance of air travel in the propagation of the virus (32).

The traditional SIR (33) (susceptible, infected, recovered) model can be recovered from our allometric equation by setting b = 1. An area of further study is to understand how some illnesses are better described by the classical SIR approach. In order to formulate the allometric model we hypothesized that the behavior of healthy populations follows scale-free dynamics. This allometric property could arise from small world networks (34) in human society. Since COVID-19 seems to have a significant component of asymptomatic transmission, the propagation of the virus has access to the two important features of small world networks: a clustering coefficient larger than in random networks, and a logarithmically increasing diameter that depends on the number of interactions. In this context, when a virus propagates symptomatically, then we assume that individuals reduce their interactions, particularly those that go across clusters, thus not allowing the virus to propagate across the network and only interacting with their nearest connections. However, further modeling and analytical work is required to test these ideas.

Our analysis confirms that the power-law dynamics of COVID-19 is correlated to human dynamics. In fact, *α* is correlated to air travel passengers per country. In the US, *α* is also correlated to air passengers per state and metropolitan area. Furthermore, *α* is correlated with some types of vehicle travel. Our simplified allometric model was able to predict the evolution of the illness several weeks in advance. These observations let us to conclude that current models of disease transmission should integrate power-law dynamics as part of the modeling strategy and not only as an emergent phenomenological property.

## Methods

### Data

The data for the number of cases of COVID-19 across the World was obtained from the Github repository of the Johns Hopkins Center for Systems Science and Engineering (35). The data file contained information for sub-regions in Australia, Canada, and China (36).

The data for US states (us-states.csv) and counties (us-counties.csv) were obtained from the New York Times database(37).

The data for World air travel was obtained from the World Bank (38). We used the data reported for 2019. The data for Norway and Sweden was obtained from www.statista.com (39, 40).

The data for the number of air travel passengers for the US was obtained from the 2019 airport ranking by the Bureau of Transportation Statistics (41). From the location of each airport, we computed the total number of passengers for each state. In the case of New York, we incorporated the numbers of La Guardia (located in New Jersey).

To calculate the spread of COVID-19 in the metropolitan areas served by major airports we used the US county database. We created custom software that extracted the number of cases for the counties that corresponded to a given metropolitan area. The cumulative number of COVID-19 in each metropolitan area was fitted as in all the other cases.

The data on number of vehicles miles traveled for each US state in urban, suburban, and rural areas was also obtained from the Bureau of Transportation statistics that corresponded to 2017 (42).

The data on the 1918 pandemic in Philadelphia and the Ebola pandemic in West Africa were obtained from Fig 1 and Fig 2 of (25) by data extraction using the software ‘WebPlot Digitizer’ (43).

### Model

We solved the allometric growth equation (Eq. 3) where *b* < 1 and *γ* and *β* are constants. If *γ* is much larger than *β*, then the growth of *I*(*t*) saturates to reach the equilibrium value:

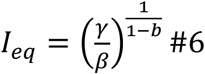

If we make a change of variables in Eq. 3:

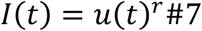

we obtain,

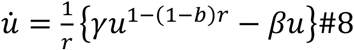

We set 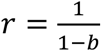 which turns Eq. 8 into:

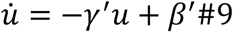

with

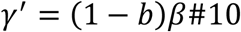

and

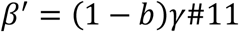

Thus, we get

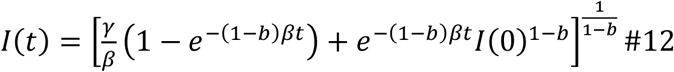

where *I*(0) is the number of cumulative cases at *t* = 0. In the case for a very extended time 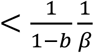, the intermediate time regime is described by:

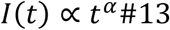

with

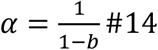

We used Eq. 12 to fit the positive cases of COVID-19 throughout this study. We used a Log-Log transformation of the data to fit the power-law function, and a Log-Linear, for the exponential. In this way, we fitted a line for the power-law and exponential cases. For the allometric function, we did not perform any modifications. In all instances we used the *fit()* function in Matlab with a non-linear least square algorithm. Individual fits are reported with their 95 % confidence intervals (CI95). Pooled data is reported with standard error of the mean (SEM). All scripts and data files are available at www.github.com/Santamarialab.

## Supporting information

Australia

Canada

China

World

## Data Availability

All data was collected from publicly available databases. All analysis scripts are available upon request.

https://github.com/SantamariaLab/COVID-19-Allometric-model

