## Supplementary Material for "The allometric propagation of COVID-19 is explained by human travel"

The Excel spreadsheet files contain tables with the results of the curve fits. The columns for **alpha** refer to the power-law exponent. The columns **b**, **beta**, and **gamma**, correspond to the allometric fits. The **tau** column is derived from the exponential fit. The **BestFit** column indicates which fit was best. The **AllRpos** is an ordinate that refers to the original data structure in the Matlab files. The **Date** column is the date when analysis started. The **AirPass** column is the number (in 1000s) of air passengers in 2017.

- Wolrdanalysis.xlxs

We extracted data for subregions in several countries. The tables have the same structure as the World Table without the air passengers. Files:

- Australiaanalysis.xlsx
- Canadaanalysis.xlxs
- Chinaanalysis.xlsx

The US table has the same structure as the World table plus columns for the average number of **urban**, **suburban**, and **rural** miles traveled by state. Files

- USanalysis.xlxs

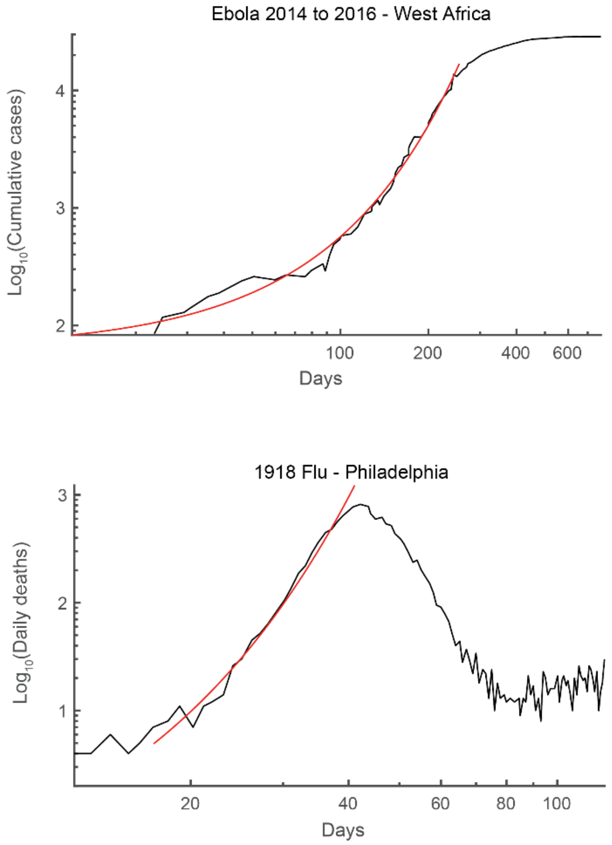


Figure S1. (Top) Total positive cases during the Ebola epidemic in West Africa during 03/01/2014-05/01/2016. The red curve shows the exponential fit ($e^{0.0218t}$). (Bottom) Weekly influenza deaths during the 1918 pandemic in Philadelphia during 09/01/1918-01/01/1919. The red curve shows the exponential fit ($e^{0.228t}$).

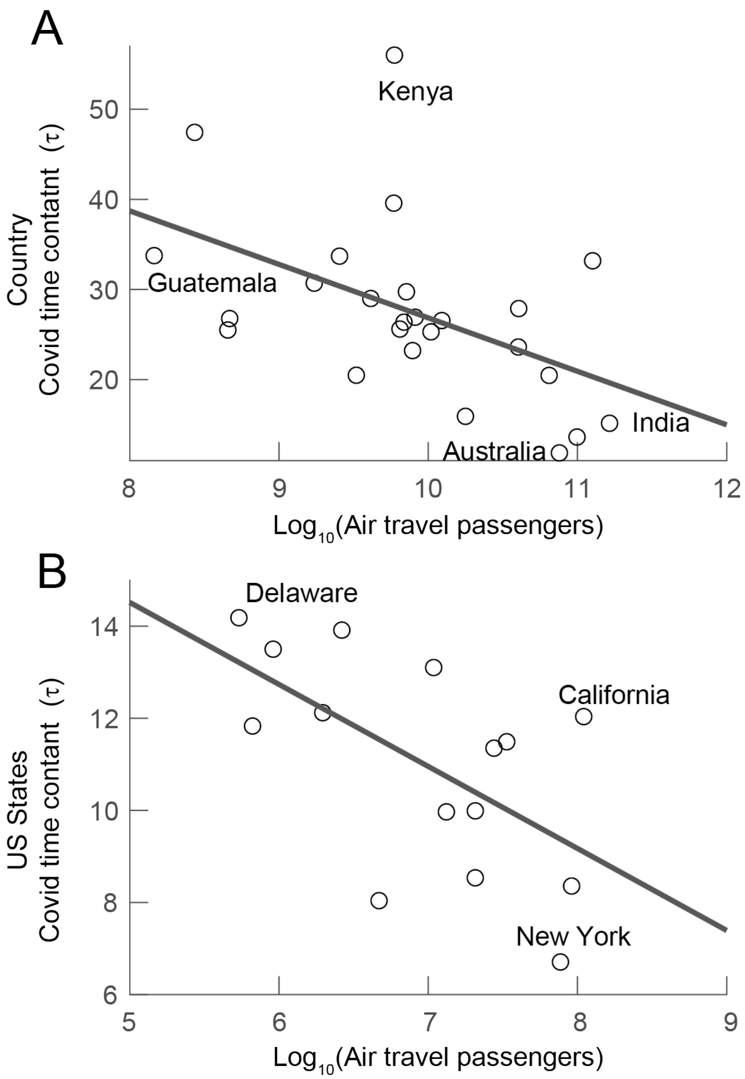


Figure S2. The time constant of the spread of the cumulative cases of COVID-19 correlate with the scale of air travel. A) The time constants obtained from fitting an exponential function to the data for the World. Only those countries that shows a better fit than the power-law were analyzed. B) The same for the amount of air travel in US states.

Table of correlations and p-values for the magnitude of air passengers and vehicle travelled miles modalities per US state. Figure 5 in the main text:

| | $Log_{10}$(Air Passengers) | |
|---|---|---|
| | r | p-value |
| Urban | 0.34 | 0.05 |
| Suburban | 0.16 | 0.37 |
| Rural | 0.10 | 0.57 |
